# Investigating the Usability, Feasibility, and Effect of a Virtual Reality Cognitive Training System on Brain Cancer Patients with Mild Cognitive Impairment: A Quasi-Experimental (Single-Arm) Pilot Study

**DOI:** 10.64898/2026.05.18.26353031

**Authors:** Anthony Faiola, John Villano, Saira Soroya

## Abstract

**(1) Background:** Brain cancer is the ninth leading cause of cancer death in the US, with approximately 76,000 newly diagnosed cases annually. Studies show that at time of diagnosis, up to six-months post-treatment, 50%–80% of brain cancer survivors (BCS) report cognitive dysfunction. Mild cognitive impairment (MCI) has gained increasing attention as a persistent disability experienced by up to 75% of all BCS, which affects memory, concentration, executive function, etc. Studies show cognitive training with computerized gaming as improving cognitive function for patients with stroke, dementia, and Parkinson’s. It is of significant clinical interest to develop innovative interventions that reduce MCI. Aim: To improve cognitive performance of BCS suffering with MCI by evaluating the feasibility, acceptability and effect of a Virtual Reality Cognitive Rehabilitation Training (VR-CRT) platform during four weeks of cognitive training. **(2) Methods:** We employed a quasi-experimental pretest/posttest non-randomized/non-blinded single-arm design for 4 weeks, with an experimental group (n=6, after attrition) using VR-CRT. Participants were selected based on convenience sampling using the electronic medical record to identify qualified patients, guided by inclusion/exclusion criteria. Feasibility was defined by retention as >80%, with usability testing using the System Usability Scale (SUS) and NASA-TLX surveys. The Hopkins Verbal Learning Test (HVLT), Controlled Oral Word Association (COWA) test, and Trail Making A-B (TM-A/B) test were used to measure cognitive performance, comparing baseline to post week-four. **(3) Results:** The feasibility criteria of >80% was met. All SUS and NASA scores were in the higher index, suggesting a high degree of usability, with low workload demand. For effect, the COWA findings showed a significant improvement (41.38%), with a paired sample T-Test confirming that the participants’ COWA scores improved significantly from pre- to post-intervention (*p* = 0.03), indicating enhanced verbal fluency and executive functioning after intervention. HVLT (combined) showed improvements of 18.75% for Form A and 11.32% for Form B, which also showed a significant improvement (*p* = .04) in the retention discrimination index from pre- to post-test. The TM-A/B test showed an improvement (25.97%), suggesting that the participants spent less time completing both parts A and B, but was not statistically significant. **(4) Conclusion:** This study fulfilled our aim to demonstrate modest to significant cognitive improvement using VR-CRT with brain cancer patients with MCI. Despite the small sample size, we believe the use of virtual reality will lead to important advances for patients with MCI, particularly the frontal lobe brain region, expressed in executive function.

## 1. Introduction

Brain cancer (glioblastoma) is the ninth leading cause of cancer death in the United States, with approximately 76,000 new cases annually [1,2]. Brain cancer survival rates are 12%, and 18,760 deaths were estimated to occur in 2024 [3]. Studies show that at the time of diagnosis (including 6 months following radiation and chemotherapy), 50%–80% of brain cancer patients report cognitive dysfunction, with some data showing cognitive impairment as high as 91% [4]. Within the glioma population, cognitive impairment ranks first among all concerns following treatment [5],[6]. Severity of impairment varies. In many cases only the patient and close family members notice changes, which can be minor to severe and can disrupt daily routines, jobs, and social functioning and can result in lifelong disabilities [7],[8].

Specifically, cancer-related cognitive impairment (CRCI) is prevalent in up to 75% of all cancer survivors [9,10],[11]. Statistical estimates suggest that by 2020 there were over 70 million cancer survivors (all types) worldwide, which highlights the global impact of CRCI and why it is a significant concern for many health care professionals [12, 13],[14],[15]. As a persistent cognitive disability frequently experienced during or after treatment [16],[10], CRCI has gained increasing recognition as a leading issue for BCS [17]. One report states that 35% of cancer survivors continue to exhibit CRCI for months and years following treatment [18]. Studies confirm that brain function networks are disrupted in posttreatment brain cancer patients [19],[20, 21]. Within this pathophysiological framework, acute and chronic cognitive impairment result from widespread neural network dysfunction and disruption, cortical network dysconnectivity, and neurotransmitter imbalances associated with acute and chronic widespread brain injury, such as with neurotoxicity. Affected areas include all cerebral lobes as well as subcortical areas, which researchers suggest impact patients’ large-scale distributed neuro-networks [19],[22],[23],[24].

The devastating effects of CRCI[25],[26],[9],[27],[28] affect working memory, attention, processing speed, and executive function as well as the disabling psychosocial effects of depression, anxiety, deterioration of social and family relations and work life [17],[10],[1]. Consequently, CRCI has a destructive impact on health-related quality of life [8],[10],[29].

Great clinical and scientific effort has been applied to reduce cognitive impairment for both BCS and other cancer survivors with mixed results. Such efforts have included physical exercise [30],[31], mindfulness training [32], and crossword puzzles[33, 34] as well as forms of digital cognitive stimulation [35],[36],[37],[38]. Pharmacotherapeutic interventions have helped manage some cognitive symptoms but often with conflicting results and serious side-effects as well as no standard of long-term care being established for widespread application [39],[40].

Early studies have shown how cognitive rehabilitation using computerized games has improved cognition [41–44], specifically, attention and working memory for patients with stroke [45],[46] dementia [47], elderly with Parkinson disease and other cognitive impairment-related disabilities [48–50],[51] and childhood brain tumors [52]. The lack of interventional treatment to improve CRCI for cancer survivors is well documented, especially cognitive rehabilitation services that offer scalability and reach [53],[54],[55]. Besides the unavailability of cognitive therapy support in outpatient clinics, existing interventions are difficult to find and set up in routine/supportive care and are usually not adapted to patients’ schedules and cognitive needs. Studies show low adherence to and compliance with the intervention [56]. Increasingly, the use of digital solutions such as computerized gaming has proven to be effective in the improvement of brain health [57].

Despite the high rates of cognitive disability, a majority of BCS do not receive effective posttreatment brain care [58, 59]. Virtual reality (VR), as a new form of digital intervention, has the advanced scientific potential for posttreatment cognitive therapy because of its deep, direct interaction with sensory and attentional networks [60–66].

VR produces cognitive stimuli that engage the frontoparietal network (involved in visuospatial-processing) and the salience network necessary for selective attention.[67] Through synergistic effects, VR promotes activity of the locus coeruleus-norepinephrine system,[68, 69] while offering a powerful effect on the regulation of multiple memory systems, e.g., the hippocampus is responsible for tagging memories with respect to context (time/place) [70], functioning as a reciprocal connection to the rest of the brain through the pathways of the neo/frontal cortex and the basal ganglia.

In addition to allowing for reproducible, cognitive processes underlying attention, memory, information processing, logical sequencing, and problem-solving, immersive VR experiences may aid in the stimulation of time and place neurons, i.e., in the organization of events in different relational dimensions that provide cognitive maps that form episodic memory [71],[72].

Contrary to paper-based crossword puzzles or digital 2D tablet-displays, VR immersive cognitive stimuli produce greater cognitive engagement and pathway integration whenever players begin to understand their global frame of reference, i.e., within the immersive environment through which they are navigating [73]. Moreover, VR immersive environments are highly interactive, activating the visual, vestibular [74], and proprioceptive systems [75]during the execution of a task in immersive spaces. With this, the VR experience for BCS has the potential to increase processes that result in a number of receptors releasing neurotransmitters and from this the potential for neurogenesis [76] and increased plasticity, as previously observed in stoke patients [77],[78], brain injury patients [79],[80], Alzheimer’s disease [81], and non-patient studies [82].

It is of significant clinical interest to brain cancer survivorship to develop a novel non-invasive approach to improve cognitive function after chemotherapy, radiation, and other oncology treatments. Chemotherapy particularly modulates neurogenesis and gliogenesis, that is, the forming of non-neuronal glia populations from neuronal cells, which result in impaired neuronal impulse conductions. Innovative technologies, such as the proposed VR gaming platform, has the potential to improve post-treatment cognitive health, while mitigating increased downstream cognitive impairment, as well as improving mental health, i.e., lowering anxiety and depression and the overall health-related quality of life.

Our proposed VR platform, referred to as: Virtual Reality—Cognitive Rehabilitation Training (VR-CRT), could bring significant scalable innovation for the survivorship community in brain function recovery. VR-CRT is intended to be delivered at clinical sites, but used in patient homes, while researchers monitor remotely. The VR-CRT platform provides privacy, convenience, flexibility, and accessibility, with high potential for scalability and reach. Backend scores and execution time tracking highly promote ongoing engagement and insight into participant progress.

VR-CRT offers advantages over traditional computerized cognitive training programs due to its immersive 3D VR environment with embedded selection addition exercises. Our VR training platform is intended to engage brain cancer survivors’ cognitive function, such as attention, working memory, executive function, and visuospatial memory, offering a unique mechanism of neurotransmitter stimulation. Moreover, VR-CRT offers a scenario goal-driven framework that rewards “players” with points, scoreboards, and feedback at the end of each module to promote engagement and incentivize use. Automated game score tracking encourages aspirational behavior and goal setting with each module completion. VR-CRT was designed with three interconnecting game levels, each with their own unique navigable (progressively demanding) brain stimuli, along with the ability for new training module integration (plug-ins) to keep the game new and challenging to players.

Our study addresses a significant knowledge gap [66] in health VR research related to interventional dosing, duration and timing, and acceptability for brain cancer survivors with CRCI. Methodological limitations in prior studies include heterogeneity in intervention design, lack of active comparators and blinded assessments, differences in amount of training, and brief follow-up periods [83]. As next steps, a rigorously conducted control pilot trial is needed that measures cognitive performance among BCS. In our study we investigated the feasibility and acceptability and estimated effect of a novel (already developed) VR game platform versus (paper-based) word-search puzzles (WSP). The results of our study provides valuable data with which to conduct an adequately powered efficacy trial.

As such, we posit that our proposed method would increase the signaling pathways of the brain, with the goal of restoring those regions of the brain that contribute to increased neurogenesis and synaptogenesis, which would result in improved cognitive performance [84–87]. The goal of this research is to validate the degree to which the VR-CRT intervention could improve the cognitive performance of brain cancer survivors suffering from CRCI, with a focus on memory, attention/focus, processing speed, and executive function.

As such, we hypothesized that (1) (non-BCS) participants (*n*=10) conveniently assigned to the usability testing of VR-CRT would show scores that reflect high usability and low cognitive demand and (2) BCS participants (*n*=6) using VR-CRT would demonstrate that a larger study was feasible and that after using VR-CRT for four weeks, cognitive scores would trend toward a higher (improved) total index from baseline.

## 2. Methods

### 2.1. The Intervention

VR-CRT was developed 2021-24 and consists of a suite of 18 modules divided into three levels of increasing cognitive demand. Cognitive training categories focus on attention, memory, processing speed, and visuospatial memory. VR-CRT uses 3D immersion embedded with selective attention (SA) exercises to improve time-order judgment, visual discrimination, spatial-match and coordination, instruction-following, and memory. It also challenges cognitive time management by incentivizing for reward/non-reward and encourages strategic thinking. The game also summarizes progress at the end of each level, and advances participants through more challenging modules to build mastery. The participant navigates VR-CRT using hand-held controllers. Visual cues and audio instructions guide the “player” towards the “mission” target to complete each level, e.g., reaching a goal destination, completing a matching exercise, identifying the correct object, performing memory tasks.

The game begins with the patient avatar being spoken to by a physician, informing them of their health status and directing them to leave the hospital, from the patient gets out of bed and begins to navigate out of their hospital room. As the game progresses, these “goals” become more challenging with attentional targets, increased distractors and faster-moving objects. Each SA exercise increases in the speed of objects moving, number of objects, complexity of the subject of the images, and multi-layering of sounds, all combined to cause additional cognitive demand and recruitment of increased attention, with the intent to engage memory, concentration, and processing speed [88],[89]. VR-CRT captures detailed backend participant-level performance statistics which provide additional game performance measures. Participants receive positive reinforcement via scoreboards, sounds of applause, and large encouraging visual messages. See Figure 1 for examples of VR-CRT game scenes and selective attention exercises.

**Figure 1.**
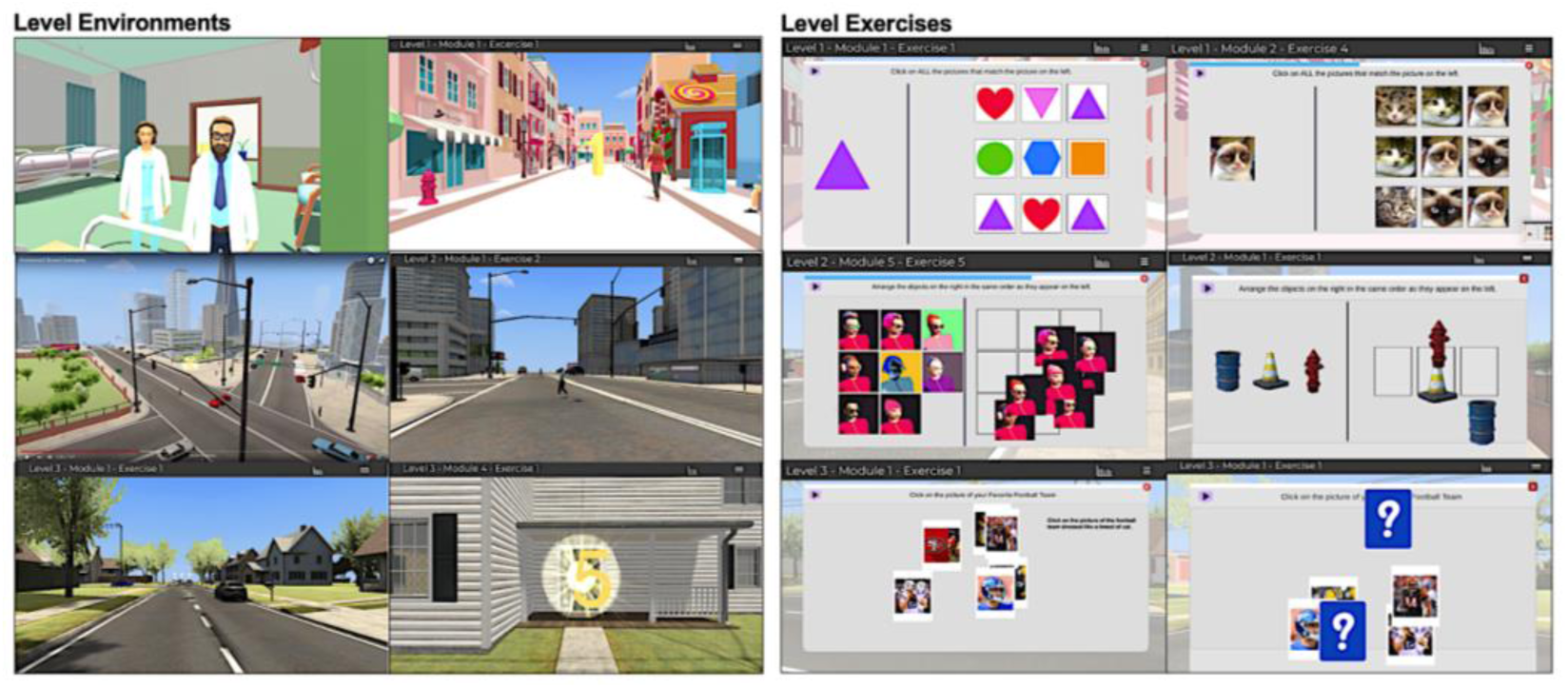
Examples of VR-CRT game interfaces of six environments and six exercises.

VR-CRT was also designed with a backend tracking system to automatically capture all user activities. Data collected include: 1) Number and duration of sessions per participant (how many times they logged in), and 2) Activity characteristics per participant, i.e., session date/time, highest game levels achieved, number of errors and attempts, and time needed to complete modules. While the VR-CRT game is accessable through the Meta store, the web-based platform GitHub hosts the data repositories for storing and managing code and player tracking. Simultaneously, Microsoft Firebase provided backend services to build and manage the VR-CRT real-time database, user authentication, and analytics, allowing seamless multiplayer experiences and efficient data management.

Personal health information (PHI) was not recorded or collected during the game play process i.e., there was no physical or mental health measures collected. The backend of the system identified objective activity measures in the execution of the game, providing general insight into changes in participant performance, but was hardly usable to validate any improvement in cognitive function.

Regarding HIPPA compliance and Personal Health Information (PHI) safeguards, the VR-CRT platform uses encrypted code on a secure server when data is collected. However, it should be noted that no study participant or patient personal or identifiable health information is in anyway input into the system during registration or throughout the study process. A randomized patient code is used to connect the smartphone to the bedside tablet in the cloud server with no connection or affiliation to the patient or any family member. This configuration was intentionally devised to avoid any issues related to HIPPA policy.

### 2.2. Participants

#### Usability Participants

The participant population for the VR-CRT usability study was recruited from the University of Kentucky (UK) general academic community. Inclusion and exclusion criteria included any staff, faculty, and students from UK who was willing to participate in the study. See Table 1 for inclusion and exclusion criteria. Usability participants consisted of ten (*n=10*) participants, two males and eight females (20%/80%).

**Table 1.** Eligibility Criteria for the VR-CRT study.

| << Inclusion Criteria >> |
| --- |
| • Brain cancer survivor, aged >18 years and English speaking.<br>• >30 days beyond completed cancer treatment, chemotherapy, radiation, and surgery, but may still be on target pharmaceuticals.<br>• Physically, cognitively, and emotionally able to wear and operate a VR game headset and hand-controllers.<br>• Able to provide written consent.<br>• Available to use the target intervention (at home) for 6 times per week for 4 consecutive weeks, for 15 min per day, adding on 5 min per week.<br>• Own a cell phone by which research staff can contact them. |
| << Exclusion Criteria >> |
| • History of acute neurologic injury such as stroke, traumatic brain injury, intracranial hemorrhage, or seizures.<br>• History of mental illness and other neurodegenerative diseases or severe psychiatric illness which is not well managed as determined by the patient’s electronic medical record (EMR).<br>• Severe, uncorrected vision or hearing impairment that prevents participation and normally engaging the intervention or outcomes assessments; as determined by the patient’s EMR.<br>• Chronic cognitive or physical function deficit preventing participation in study interventions or assessments as determined by the patient’s EMR.<br>• Uncorrected hearing or vision impairment including legal blindness.<br>• Suspected or confirmed drug use, intoxication, and/or overdose.<br>• History of severe vertigo, motion-induced sickness, headaches, light sensitivity, seizures, or prior intolerance of virtual reality (VR).<br>• Incarcerated or patients unable to provide consent. |

*2.6. Recruitment and Onboarding*

Recruitment of usability participants included advertising for the study using the UK Center for Clinical and Translational Science (CCTS), Participant Rectuitment Services (PRS). Sixity IRB approved recruitment flyers were placed on CCTS PRS wall mounts, plus advertised through social media, digital monitor ads, and ResearchMatch.org ads. Within one month 34 potential participants responded to the ads via email to the principal investigator (PI). From this group 10 responded to follow-up emails and agreed to participant after a more detailed explanation of the study. Recruitment began 2-1-23 to 4-1-23. Monetary compensation was provided.

#### Experimental Participants

Although the recruitment target of this experimental arm was 10 patients under active followup care, recruitment challenges resulted in only 6 participants (*n=6*). All participants who used the VR-CRT training system were selected according to the inclusion and exclusion criteria and based on convenience sampling and suitability to the study [90]. Recruitment was executed by the physician researcher from the UK Neuro-oncology Clinic of the Markey Cancer Center (MCC). Recruitment took place following patients’ followup appointments with the physician researcher. See Table 2 for inclusion and exclusion criteria.

**Table 2.**
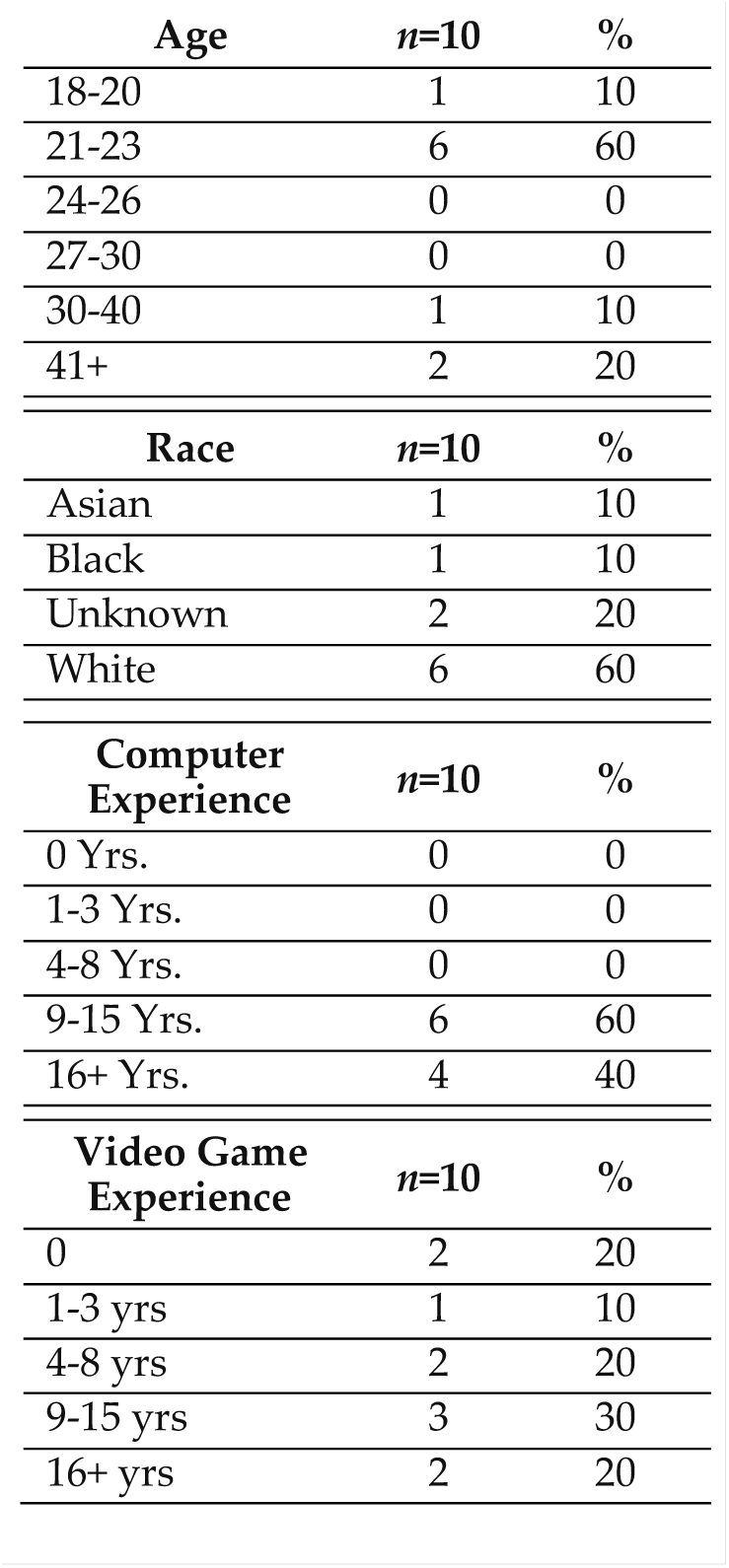
Demographic data of usability participants.

Before contacting patients, we received approval from the UK Institutional Review Board and the MCC Protocol Review and Monitoring Committee, who review and approve all cancer human subjects research, as well as all data auditing for quality control purposes. Recruitment began 11-15-23 to 8-1-24. Monetary compensation was provided to all participants.

### 2.3. Design

This study employed a quasi-experimental [91] pretest–posttest non-randomized [92], non-blinded single-arm design to evaluate the usability, feasibility, and effects of the VR system, VR-CRT, on cognitive impairment in brain cancer patients during an outpatient four-week treatment. Before testing for cognitive performance, we executed a pre-measurement usability study including two usability measures, the System Usability Scale (SUS) and the NASA-TLX survey (NASA), discussed in greater detail below.

### 2.4. Usability Study Design

Post-measurement data collection included two usability measures, the System Usability Scale (SUS) and the NASA-TLX survey (NASA). SUS was used to assess the perceived usability of the intervention, VR-CRT. It is a validated and reliable scale consisting of ten items that are rated on a 5-point Likert scale ranging from 0 to 4 [93]. The SUS quantifies the usability of products and services, including software, mobile apps, websites, or any interactive device with an interface. Two examples of questions include: “Thought the system was easy to use,” “Found the various functions in this system were well integrated,” and “Thought there was too much inconsistency in this system.” User response options range from 1 (Strongly Disagree) to 5 (Strongly Agree). Researchers used data from 241 SUS usability studies to create a curved grading scale, from which they found that the mean SUS score was 68, with 50% falling below and above it. A mean SUS score > 80 could be considered “good” as evidence of an above average user experience, < 70 could be considered as having usability issues As such, a top and bottom 15 percentiles correspond to A and F grades, with further subdivisions for A+, A, A-, and additional breakdowns for grades of B and C [94],[95].

The NASA survey is a validated instrument designed for participants to assess the subjective workload of their use of the VR-CRT [96],[97]. Workload of a task depends on a variety of factors such as the nature and difficulty of the task as well as the aptitude and attitude of the individual. NASA consists of two parts divided into six subjective subscales that are represented on a single page, including: Mental Demand, Physical Demand, Temporal Demand, Performance, Effort, and Frustration. Each NASA subscale is scored from 0 to 100 in 5-point increments. While there is no standard NASA baseline, studies suggest that low workload are scores below 40, moderate workload scores are between 40 and 70, and high workload scores are above 70. Higher scores in each subscale indicate higher demand of the task, and in some cases, a lack of success in executing the task. Scores from all subscales were averaged for the total unweighted NASA score.

### 2.5. Experimental Study Design

The study design, interventional dosing, and all three baseline cognitive measures parallel this proposed study. The measures include the: Hopkins Verbal Learning Test (HVLT), Controlled Oral Word Association (COWA) test, and Trail Making A-B (TM-A/B). We selected participants based on convenience sampling and suitability for the study [71], and we initiated the study in the Neuro-oncology Outpatient Clinic of the Markey Cancer Center (MCC) at the University of Kentucky and completed the study in the homes of the participants. See Table 1 for a list of inclusion and exclusion criteria.

### 2.6. Recruitment and Onboarding

The 12-month MCC brain cancer patient surveillance population is approximately 130 per year, which was sufficient to support the recruitment of 10 brain cancer survivors (*n*=10 Experimental) after inclusion and exclusion criteria. A 30-40% (or higher) attrition rate was anticipated with a final sample of approximately 10 patients (*n*=10 Experimental). Our recruitment period was from January to June 2024. Brain cancer patients were screened for cognitive dysfunction at the MCC outpatient clinic by the neuro-oncology director and research team. Once identified, study personnel contacted eligible survivors who are 30 plus days post-treatment for consent and enrollment. Due to severe illness of two patients and challenges in recruitment, only 6 patients completed the study.

Following consent, participants began the onboarding phase, beginning with 3 baseline cognitive performance tests, discussed below. After their cognitive baseline assessment, participants were trained how to use the VR headset and training VR-CRT software. Training sessions were in a seated position. All training sessions were delivered with participants in a seated position, at which time we observed for motion sickness symptoms or any other mental or physical challenges, complaints, or conditions. Participants were asked to self-report if they had a history of motion sickness.

Experimental dosing was 6 days per week for 4 weeks, starting at 15 min (Wk. 1) and increasing 5 min per week. This achieved 540 cumulative minutes of cognitive training, which aligns with duration associated with benefit in prior literature [98]. Participants received text and/or phone call reminders or general check-in calls to assess if they were maintaining their dosing schedule or have any difficulties using the intervention.

The experimental participants received the hardware and software free of charge, but with a sign-out consent to confirm their agreement to return the equipment once the study was completed. Off-the-shelf VR equipment (Meta Quest 2/128 GB with WIFI) was used for the delivery of the VR-CRT intervention. VR-CRT was already loaded in the headset, with pre-assigned login setup. In addition to the encrypted VR-CRT platform for for ensuring PHI security, as noted above, participant login codes were anonymous, with connection to the patient’s name or patient ID number. In this way patient identify was protect, thereby avoiding any issues related HIPPA policy.

Participants were given detailed verbal instructions and safety education on the VR technology. The training process took place in the neuro-oncology outpatient clinic or a public library private reading room near their home, whichever was more convienent for the participant. In the latter case (library) onboarding locations was usually the most successful due to their convenience, with no difficulties, incidences, or safety issues reported by the patient.

### 2.7. Treatment Dose and Schedule

Participants were provided a schedule of VR-CRT dosing. See Figure 2 for the instructions and daily intervention dosing schedule that participants received during their onboarding. The schedule covers the day-by-day dosing routine for 4 weeks. Once onboarding was complete, participants were sent home with the VR equipment. Once at home, participants logged into VR-CRT with their designated (anonymous) user login name. Because VR-CRT was developed to capture all participant activity in the cloud all sessions will be monitored remotely. This enabled researchers to observe participant progress and general activity in real time. We also monitored the participants more regularly via phone to assess their progress. No in-home visits were scheduled but the team provided daily support as needed for troubleshooting.

**Figure 2.**
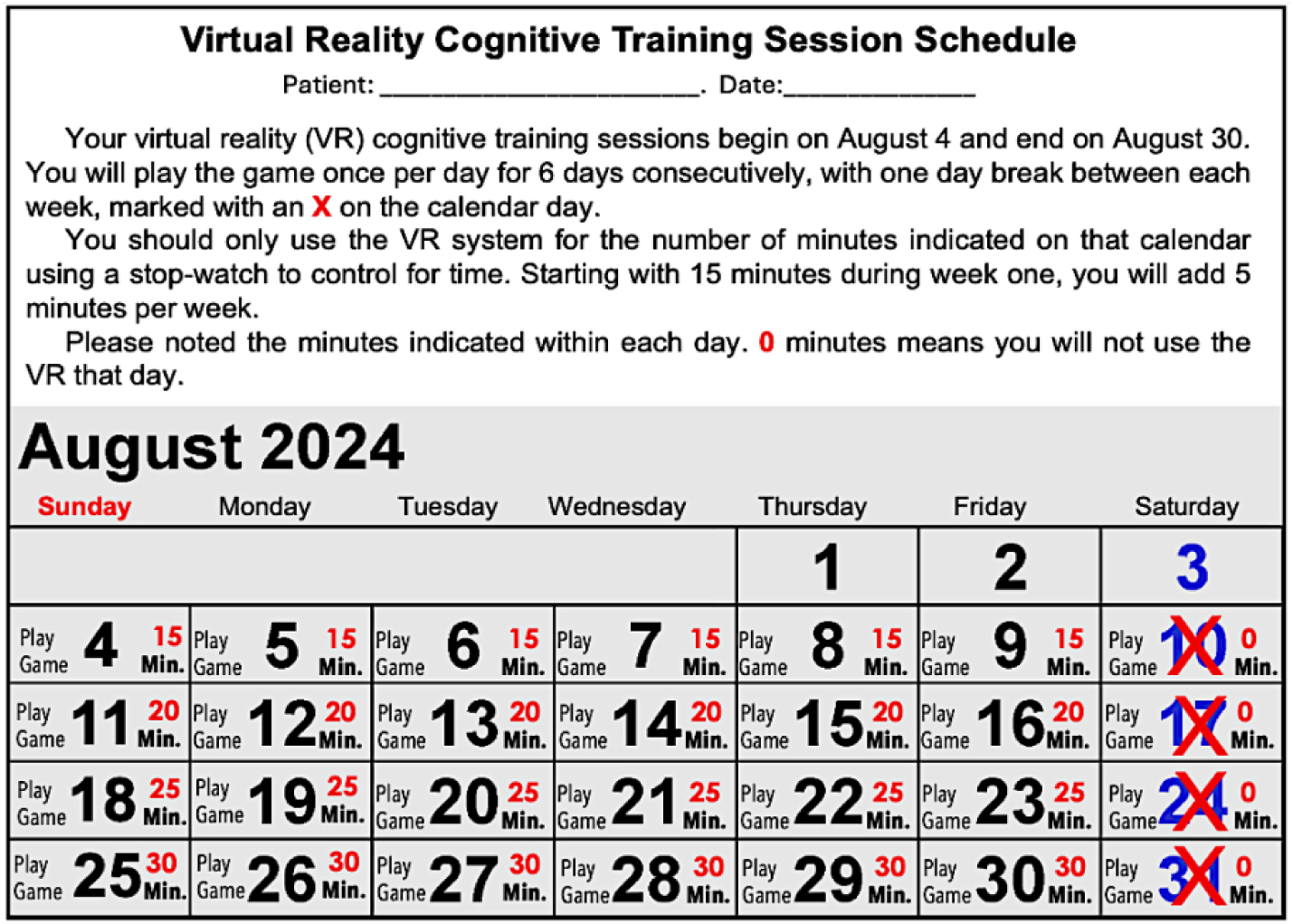
Examples of participant VR-CRT gaming schedule.

### 2.8. Cognitive Effect Measures

Cognitive performance scores were obtained using the Hopkins Verbal Learning/Memory (HVLM), Controlled Oral Word Association (COWA), and Trail Making A-B (TMa/b). Data collection provided scores at baseline (pre-training) and post-intervention (4 weeks later). Intervention effect sizes were derived from the estimated mean and standard deviation of the difference of change in scores from baseline to end of intervention [99]. All assessments were performed at the Markey Cancer Center (MCC) inpatient neuro-oncology clinic or at a public library reserved reading room location near the participants’ home. VR-CRT’s backend data collection also captured real-time gameplay data to assess processing speed and effects on the functional response times to complete tasks in over 80 selective attention exercises.

### 2.9. Feasibility

The feasibility of our study depended on patient willingness to participate, given their health condition, availability, and general interest in contributing to scientific discovery [72]. Criteria for determining feasibility for cancer patients (in general) include recruitment rates and retention rates [73] at around >70%. However, due to the severity of brain cancer, we (a priori) defined retention as >60% (>6 participants out of 10) able to complete the 4-week experimental arm [100],[101]. Retention is related to the frequency of participants’ completing the study, including the proportion of dropouts throughout the four weeks of data collection. The retention rate was calculated as the number of patients completing all four weeks of the study. The research team recorded all adverse events occurring throughout the four-week study.

#### Data Analysis

Due to the small sample size and experimental nature of this pilot study, descriptive statistics were the primary means of data analysis for both the usability testing data and the experimental data. However, given the small sample size, non-parametric statistical tests were used to examine differences (if any) across the three measures: HVLT, COWA and TM-A/B. The Wilcoxon signed-rank test was applied as an alternative to the paired-samples t-test.

## 3. Results

### 3.1. Usability Findings

#### 3.1.1 Demographic Findings

For usability participants, we collected demographic data, including age, race, computer experience, and video game experience. See Table 2. For experimental participants, we collected demographic data, including gender, age, and race. Given that this was a pilot study, we decided not to collect participant data related to education, annual income, current or prior mental health, or level of mobile app or technology acceptance, as might be measured using the Technology Acceptance Model [102].

#### 3.1.2 System Usability Scale Findings

SUS scores were analyzed in two ways. Raw score ratings are shown in Table 4 for all participant family members per each question, with an overall mean score of 4.20 (SD = 0.84). Specific SUS questions are listed in Table 4. SUS scores were also analyzed to determine an overall satisfaction score according to original criteria method [103]. The total mean SUS score of 80 (SD = 21.02) was above average, indicating good usability with individual scores ranging from 57.5 to 100. Table 5. Results of the System Usability Scale questionnaire. Total score range: 0–5, including to the collective (n=7) mean and standard deviation per each question.

**Table 3.** Demographic data of experimental participants.

| Gender | n=6 | % |
| --- | --- | --- |
| Male | 4 | 66.67% |
| Female | 2 | 33.33% |
| Age | n=6 | % |
| 30-39 yrs. | 2 | 33.33% |
| 40-49 yrs. | 3 | 50.00% |
| 50-59 yrs. | 1 | 16.67% |
| Race | n=6 | % |
| White | 5 | 83.33% |
| Hispanic/Latino | 1 | 16.67% |

**Table 4.** Results of raw score ratings of SUS questionnaire.

| SUS Survey Questions | Mean (SD) |
| --- | --- |
| Q1. I think that I would like to use this system frequently. | 3.86 (1.46) |
| Q2. I found the system unnecessarily complex. | 4.00 (1.53) |
| Q3. I thought the system was easy to use. | 4.43 (0.53) |
| Q4. I would need the support of a technical person to be able to use this system. | 3.71 (1.70) |
| Q5. I found that the categorized results in this system were very sensible. | 4.14 (1.21) |
| Q6. I thought that there was too much inconsistency in this system. | 4.57 (0.79) |
| Q7. I would imagine that most people could learn to use this system very quickly. | 4.43 (0.98) |
| Q8. I found the system very cumbersome to use. | 3.86 (1.21) |
| Q9. I felt very confident using the system. | 4.29 (1.25) |
| Q10. I needed to learn a lot of things before I could get going with this system | 4.71 (0.76) |
| **Final SUS Score** | **4.20 (0.84)** |

**Table 5.**
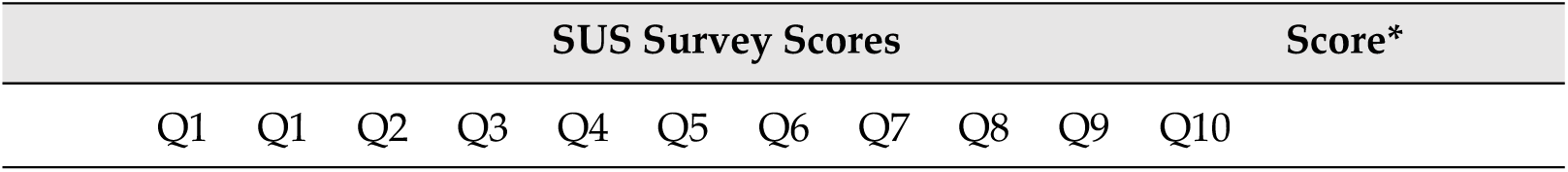
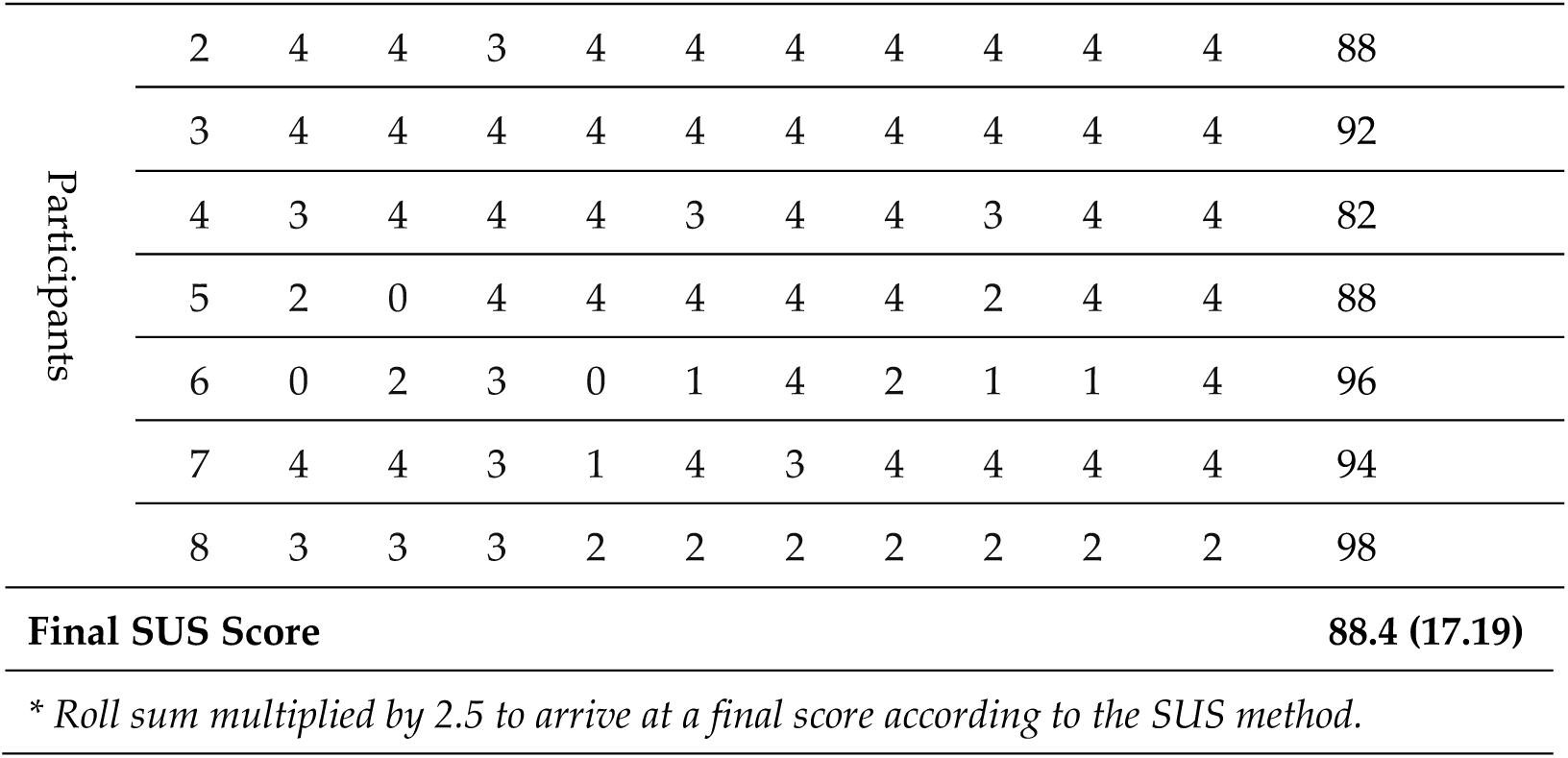
Results of the total SUS score range: 0–100, including the collective mean score per question, with standard deviation questionnaire.

#### 3.1.3 NASA Survey Findings

Regarding the the NASA survey findings, studies suggest that scores below 40 are considered low workload. All mean scores were below the threshold of 40, indicating participants’ workload experience was considerably low demand while using the mobile app, with Frustration being the lowest score (17.14) and Effort being the highest (30.00). The overall mean score was (27.50) also below 40, suggesting a low demand of task while using the intervention. Participants also reported high confidence particularly in the Frustration workload demand, and in the overall use of the app, with a standard deviation of 17.14 and 15.45, respectively. See Table 6.

**Table 6.** Results of the NASA-task load index questionnaire. Total score range: 0–100.

| Q# | Demand Type | Question | Mean (SD) |
| --- | --- | --- | --- |
| Q1. Mental |  | How mentally demanding was the task? | 27.50 (12.53) |
| Q2. Physical |  | How physically demanding was the task? | 11.00 (08.76) |
| Q3. Temporal |  | How hurried or rushed was the pace of the task? | 42.00 (22.26) |
| Q4. Performance |  | How hard did you have to work to achieve your level of performance? | 19.30 (11.76) |
| Q5. Effort |  | How successful were you in accomplishing what you were asked to do? | 27.50 (15.14) |
| Q6. Frustration |  | How insecure, discouraged, irritated, stressed, and annoyed were you? | 19.50 (13.63) |
| **Final NASA-TLX Score** |  |  | **24.47 (14.01)** |

Only seven of the nine participants that completed the study filled out the SUS and NASA surveys. However, according to usability researchers, this number of participants is more than sufficient to determine usability. [104, 105]

### 3.2. Experimental Study Findings

#### 3.2.1 Feasibility Findings

Although the recruitment process involved considerable challenges due to the life-threatening condition of the participants, we hoped for fair to good retention rates to meet the established threshold. During the 7 months of recruitment we reached out to 20 potential participants from the brain cancer population who met our inclusion and exclusion criteria, who previously were identified through convenience sampling by the neuro-oncologist team member as potentially good candidates for the study. Of these 20, 10 were Invited to participant but either said yes and changed mind soon after or did not respond to repeated phone calls. Hence, we obtained an enrollment of 50%.

Of the remaining 10 that consented, 2 participants dropped in week 2 due to increased illness and 2 participates changed their mind with concerns over their illness or general lack of interest; leaving 6 that completed the 4 weeks of the study; hence a 40% attrition rate. As such, our >60% threshold was met. Although there were several minor technical issues with the VR hardware and software, we recorded no adverse events from any of the participants in the experimental group.

#### 3.2.2 Effect Findings

Table 7 outlines all study results from the three cognitive measures. The controlled oral word association (COWA) test showed a 41.38% improvement, with the baseline total score at 33.80 and the post-intervention score at 47.80 (See Figure 3). A paired sample T-Test (Wilcoxon W) confirmed that the participants’ COWAT scores improved significantly from pre- to post-intervention (median increase from 29 to 46 words, *p* = 0.03), indicating enhanced verbal fluency and executive functioning after intervention.

**Table 7.** Three cognitive training intervention scores.

| Measures | Cognitive Function | Baseline Mean (SD) Score | Post-Intervention Mean (SD) Score | Percentage Improved |
| --- | --- | --- | --- | --- |
| Controlled Oral Word Association Test | Language ability and executive function | 33.80 (12.20) | 47.80 (18.60) | 41.38% |
| Trail Making Test A | Processing speed, attention, and task switching | 26.83 (12.04) | 25.50 ( 7.77) | 4.97% |
| Trail Making Test B |  | 56.83 (18.40) | 44.67 (17.70) | 21.41% |
| Hopkins Verbal Learning Test A | Verbal learning and recall | 18.70 ( 6.80) | 22.20 ( 7.08) | 18.75% |
| Hopkins Verbal Learning Test B |  | 8.83 ( 2.64) | 9.83 ( 1.17) | 11.32% |
| Mean % |  |  |  | 19.52% |

**Figure 3.**
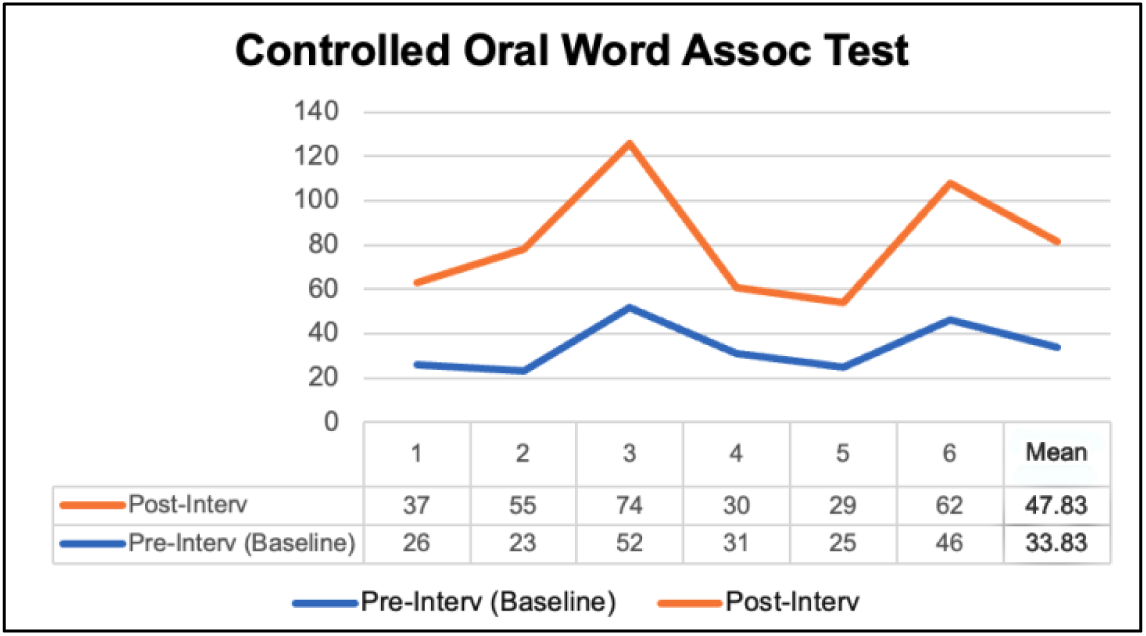
Controlled Oral Word Association test scores.

The Trail Making Test A showed a modest 4.97% improvement, while the more cognitively challenging Trail Making B showed a 21.41% improvemen,t with the baseline total score at 56.83 and the post-intervention score at 44.67. See Figure 4. In sum, participants spent less time completing both Parts A and B. However, the paired-samples test (Wilcoxon signed-rank test) indicated that these improvements were not statistically significant (P>.05). It is important to note that the small sample size (n = 6) may have limited the power to detect meaningful differences.

**Figure 4.**
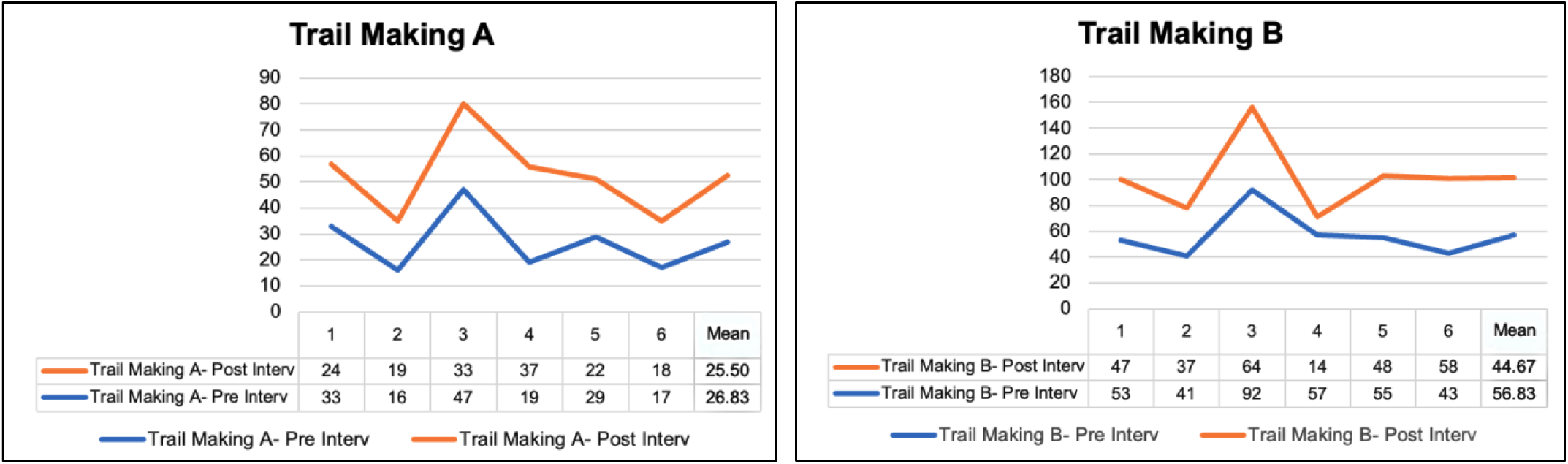
Trail Making A and B test scores.

The Hopkins Verbal Learning Test A and B (combined) showed improvements of 18.75% for Form A and 11.32% for Form B. For Form A, the mean score increased from 18.67 at baseline to 22.17 post-intervention. In contrast, Form B showed a smaller change, with the mean score rising from 8.83 at baseline to 9.83 following the intervention. See Figure 5.

**Figure 5.**
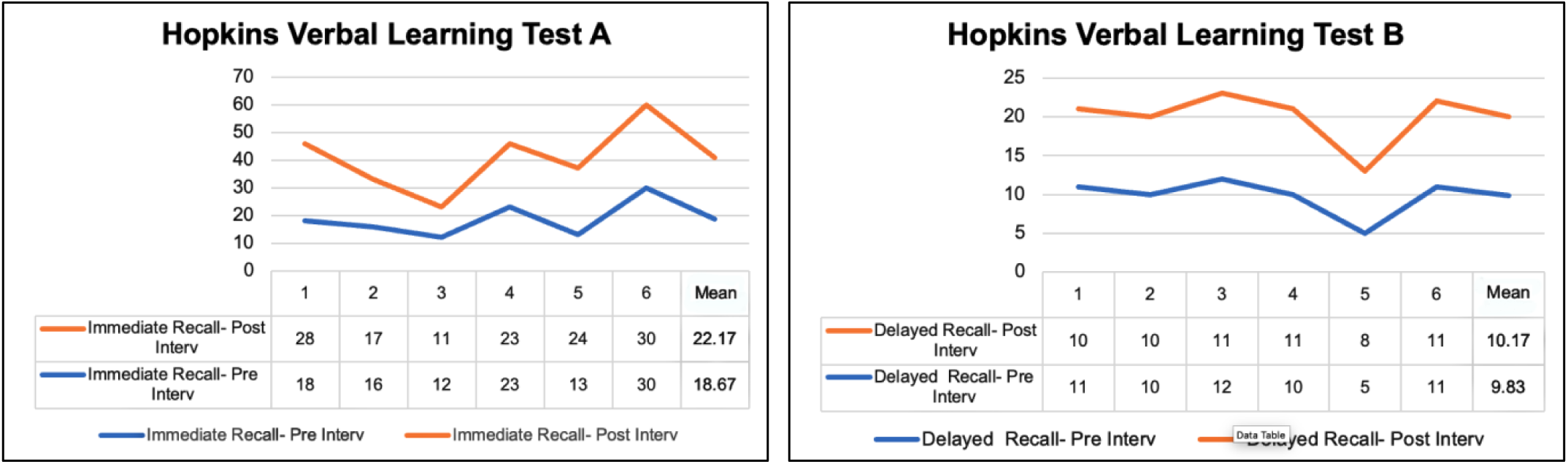
Hopkins Verbal Learning Test A and B test scores.

To verify if the improvements are significant, we ran a Wilcoxon signed-rank test that proved that a significant improvement (*p*=.04) was observed in the retention discrimination index from pre- to post-test, suggesting that participants became better at distinguishing between previously learned (target) items (M=7.17, SD=2.71) and new or irrelevant (distractor) items after the intervention (M=8.67, SD=1.97). In other words, their ability to accurately recognize correct information while rejecting false positives improved (see Table 7). In contrast, the lack of significant differences in immediate recall, delayed recall, and retention percentage indicates that the overall capacity to encode, store, and retrieve information did not substantially change over time. Participants remembered roughly the same amount of information before and after the intervention, both immediately and after a delay.

Overall, these results suggest that while the intervention may not have enhanced memory quantity (i.e., how much information is recalled), it appears to have improved memory quality—specifically, the accuracy and precision of retrieval. This pattern may indicate better cognitive control, reduced susceptibility to false memories, or improved decision-making during recognition tasks, rather than a general enhancement of memory storage or recall ability.

**Table 7.**
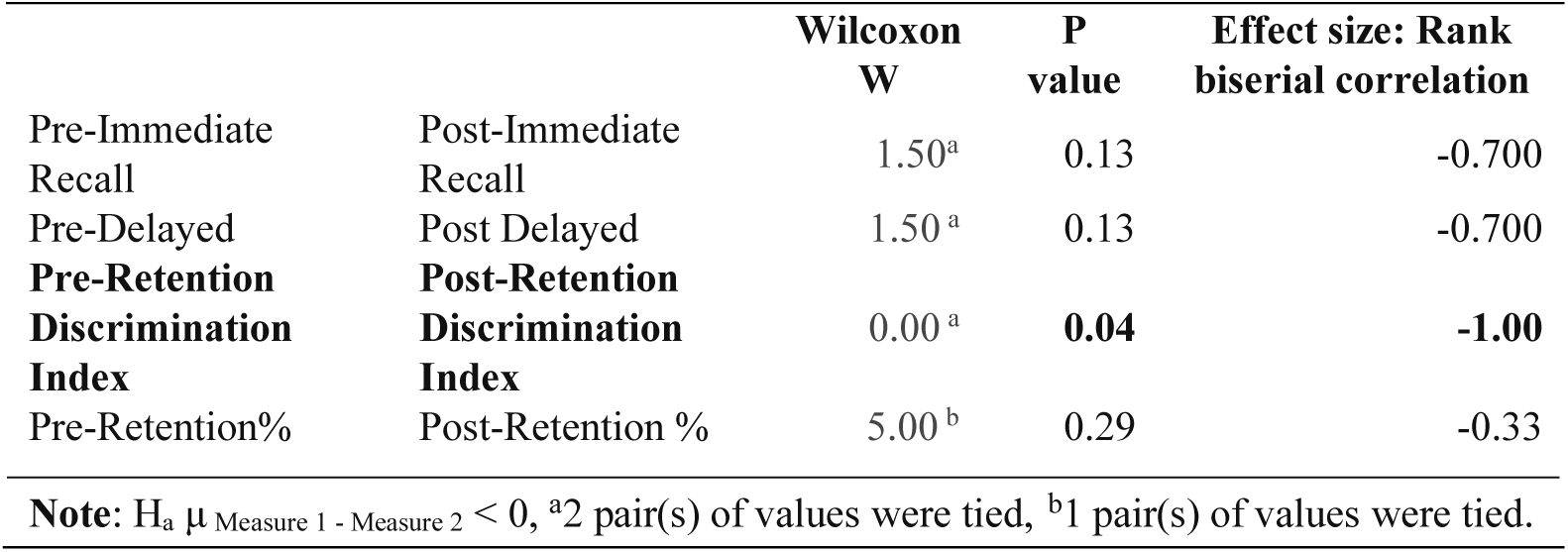
Hopkins Verbal Learning Test A and B test scores with P values.

|  |  | Wilcoxon<br>W | P<br>value | Effect size: Rank<br>biserial correlation |
| --- | --- | --- | --- | --- |
| Pre-Immediate Recall | Post-Immediate Recall | 1.50<sup>a</sup> | 0.13 | -0.700 |
| Pre-Delayed | Post Delayed | 1.50<sup>a</sup> | 0.13 | -0.700 |
| **Pre-Retention**<br>**Discrimination**<br>**Index** | **Post-Retention**<br>**Discrimination**<br>**Index** | 0.00<sup>a</sup> | **0.04** | **-1.00** |
| Pre-Retention% | Post-Retention % | 5.00<sup>b</sup> | 0.29 | -0.33 |

**Note:** $H_a \mu_{\text{Measure 1}} - \mu_{\text{Measure 2}} < 0$ , <sup>a</sup>2 pair(s) of values were tied, <sup>b</sup>1 pair(s) of values were tied.

## 4. Discussion

Based on the intended purpose of the controlled oral word association test, the 41.38% improved score (which is also statistically significant) suggests improved verbal fluency, executive function, broader cognitive plasticity, and high-order executive abilities. We posit that the reason for this improvement is due to the VR-CRT training exercises placing constant burden on the effective retrieval required for executive control over cognitive processes. Moreover, selective attention task switching required the participants to regularly shifted their attention between one task and another, with rapid adaptation to different 3D VR environments and navigational strategies. As such, cognitive shifting, as an executive function, involved conscious change in attention and visuospatial focus.

The Trail Making test directs patients to focus on visual search and motor speed skills for A and a higher degree of cognitive skill and flexibility for B. Visual search here refers to the reallocation of attentional focus from one target or location to another. According to the intended purpose of the Trail Making A and B test, the mean score difference of 26.38% reflects an improvement in visual search, processing speed, scanning, mental flexibility, and executive function. Any improvement in this test score may also be an indicator of a decrease in neurological impairment, particularly frontal lobe function. Although these improvements were not statistically significant, they suggest meaningful patterns that can inform future research on this phenomenon. The small sample size likely limited the ability to detect statistically significant changes in post-intervention scores.

HVLT is associated with verbal episodic memory, i.e., memory of everyday events and experiences related to persons, times and places. Test A and B mean score difference was 30.07%, suggesting a modest improvement in verbal learning and semantic memory, which is an indicator of a slight decrease in frontal lobe dysfunction related to executive function, related to high-order cognitive abilities, i.e., complex decision-making, problem-solving, working memory, and plasticity. Although the overall HVLT scores did not reach statistical significance, the retention discrimination index showed significant pre–post differences. This suggests that while the intervention did not produce measurable changes in global recall performance, it may have influenced more specific aspects of memory processing, such as retention accuracy or discrimination ability.

While these findings should be interpreted cautiously, (as the small sample size may have limited the power to detect change), we believe this study will lead to important advances for MCI patients. First, we hoped to observe an improvement overall, i.e., we observed for modest to above average improvement. We also expected that our original theory of the synergic effects of VR immersion and selective attention training would stimulate some neural activity, which we believe transpired. Second, we argue that it’s possible not to see improvement in all three measures simultaneously, because of the differences measuring cognitive function from diverse brain regions. While there may be some overlap in their cognitive evaluation, each has a unique focal point of analysis.

## 5. Conclusions

Beside establishing feasibility and a high degree VR-CRT usability, the aim of this study was to demonstrate modest to significant cognitive improvement using VR-CRT with brain cancer patients with MCI.

First, we hoped to observe (from this limited sample) an improvement overall, i.e., we observed for modest to above average improvement. We also expected that our original theory of the synergic effects of VR immersion and selective attention training would stimulate some neural activity, which we believe transpired. Second, we argue that it’s possible not to see improvement in all three measures simultaneously, because of the differences in measuring cognitive function from diverse brain regions. While there may be some overlap in their cognitive evaluation, each has a unique focal point of analysis. Despite the small sample of participants, we believe this study with the use of virtual reality will lead to important advances for patients with MCI, particularly the frontal lobe brain region, expressed in executive function and other high-order cognitive abilities and complex decision-making, working memory, cognitive flexibility and plasticity, reasoning, and problem-solving.

## 6. Limitations

This study has several limitations. First, the small sample size (n = 6) substantially limits statistical power and increases the risk of both Type I and Type II errors. Therefore, the significant improvement observed in the Controlled Oral Word Association Test (COWA) should be interpreted as preliminary, and the non-significant findings for the Trail Making Test and Hopkins Verbal Learning Test may reflect insufficient power rather than a true lack of effect. Conversely, it’s important to note that the majority of studies that include virtual reality technology as an intervention for both MCI and non-MCI patients, have relatively small sample sizes (15 to 60) due to the complexity and application of the technology [98],[66],[106].

Second, the study population consisted of individuals with brain cancer, which limits the generalizability of findings to other neurological populations. Although MCI may overlap across conditions, the underlying mechanisms differ, and results may not directly translate to populations such as Alzheimer’s disease and related dementias (AD/ADRD) and cancer patients suffering from the MCI-related effects of chemotherapy. Third, demographic and clinical variables (e.g., age, education, tumor characteristics, treatment history) were not analyzed due to the small sample size, which may also introduce unaccounted variability in cognitive performance outcomes.

Additionally, the absence of a control or comparison group limits causal inference, and practice effects associated with repeated administration of neuropsychological tests, which may have influenced performance, particularly in measures like COWA. Finally, other factors such as the lack of longer-term follow-up restricts conclusions about the durability of observed improvements.

## 6. Future Research

Future research should prioritize larger, adequately powered studies to validate these preliminary findings and allow for more robust statistical and subgroup analyses. This includes participants with diverse demographic and clinical profiles to improve generalizability and help identify factors influencing cognitive outcomes.

Incorporating control or comparison groups, ideally within randomized or quasi-experimental designs, will strengthen causal interpretations. Future studies may also consider strategies to minimize or account for practice effects, such as using alternate test forms of the Controlled Oral Word Association Test, Trail Making Test, and Hopkins Verbal Learning Test. Longitudinal research with extended follow-up periods is needed to assess the persistence of cognitive changes over time in brain cancer populations.

Additionally, future studies may explore whether similar intervention effects are observed in other populations with cognitive impairment, including AD/ADRD, while accounting for differences in disease pathology.

In sum, while this study yielded a small dataset without a control, our findings are promising. As such, we posit that outcomes from the proposed study will further extend these findings when applied to other MCI populations. Moreover, we posit that future AI modeling integration into VR-CRT will further enhance positive outcomes for MCI patients through expanded engagement with sensory processes, visuospatial memory, and motor behavior attentional networks, resulting in increased receptors releasing neurotransmitters [60–66].

## Data Availability

All data will be provided on request.

## Author Contributions

Conceptualization, A.F.; methodology, A.F. and S.S.; VR design, A.F.; formal analysis, A.F. and S.S.; statistical support, S.S.; resources, D.V.; writing—original draft preparation, A.F.; writing—review and editing, A.F., S.S.; and D.V. project administration, A.F., D.V.; funding acquisition, A.F. All authors have read and agreed to the published version of the manuscript.”

## Funding

This study is supported by the University of Kentucky, Center for Clinical and Translation Science, Lexington, Kentucky.

## Institutional Review Board Statement

On 10/30/2023, the Medical Institutional Review Board approved your request for modifications in your protocol entitled: A health game intervention for cancer and post-ICU patients suffering from acute cognitive impairment: A clinical study to assess a new form of brain stimulation therapy with the potential to improve synaptic plasticity. The IRB Number is 76973. University of Kentucky.

## Informed Consent Statement

Informed consent was obtained from all subjects involved in the study.

## Data Availability Statement

Data from study available on request.

## Acknowledgments

We are very appreciative of the study participants. We are grateful to Dr. John Villano, MD (College of Medicine, Division of Neuo-Oncology, Markey Cancer Center, University of Kentucky, Lexington, KY 40506) and Adria Myers, Nurse Practitioner Specialist in the Neuro-oncology outpatient clinic (University of Kentucky HealthCare), who supported oordinating patient participation.

## Conflicts of Interest

The authors declare no conflicts of interest.

## Abbreviations

The following abbreviations are used in this manuscript:

BCS: Brain cancer survivors
CCTS: Clinical and Translational Science
COWA: Controlled Oral Word Association test
CRCI: Cancer-related cognitive impairment
EMR: Electronic medical record
HVLT: Hopkins Verbal Learning Test
MCC: Markey Cancer Center
NASA: NASA-TLX survey
PHI: Personal health information
PI: Principal investigator
PRS: Participant Recruitment Services
SA: Selective attention
SUS: System Usability Scale
TM-A/B: Trail Making A-B
UK: University of Kentucky
VR: Virtual reality
VR-CRT: Virtual Reality Cognitive Rehabilitation Training
WSP: Word-search puzzles

## Disclaimer/Publisher’s Note

The statements, opinions and data contained in all publications are solely those of the individual author(s) and contributor(s).

## Notes

### Competing Interest Statement

The authors have declared no competing interest.

### Clinical Trial

NCT07596940

### Author Declarations

University of Kentucky, IRB approved this study. ID: IRB # 76973 IRB6

### Summary of Updates

None, other than I added one author that was left off by mistake in the original submission.

